# Diagnostic accuracy of image-guided fine needle aspiration cytology in diagnosis of lung tumors

**DOI:** 10.64898/2026.08.17.26360531

**Authors:** Bishwobandhu Bhandari, Mamata Tiwari, Shital Adhikari, Anuradha Khanal, Nabin Bohara Chhetri, Shikha Pandey

**Affiliations:** Department of Pathology, Chitwan Medical College, Tribhuvan University, Nepal; Department of Internal Medicine, Chitwan Medical College, Tribhuvan University, Nepal; Department of Microbiology, B.P. Koirala Institute of Health Sciences, Dharan, Nepal

**Keywords:** Fine needle aspiration cytology, Lung cancer, Biopsy, Squamous cell carcinoma, Adenocarcinoma, Small cell carcinoma, Nepal

## Abstract

**Background:** Lung cancer remains the leading cause of cancer related mortality globally. It is the second most prevalent cancer among women worldwide and ranks third among females in Nepal. Contributing factors include smoking, tobacco use, air pollution, and delayed diagnosis. Image-guided fine needle aspiration cytology(FNAC) and biopsy are widely used, rapid and diagnostic techniques for evaluating lung lesions. As FNAC is minimally invasive procedure with less complications as compared to biopsy, the goal of this study is to examine the histocytologic makeup of lung lesions and link the results.

**Materials and Methods:** This cross-sectional observational study included 65 patients irrespective of age and sex presenting with lung masses at Chitwan Medical College and Teaching Hospital during the period of eighteen months starting from April 2023 to September 2024. After clinical and radiologic evaluation, all cases underwent image-guided FNAC and biopsy. Only specimens with unequivocal malignant features were classified as positive. Histopathology served as the diagnostic reference standard.

**Results:** FNAC diagnosed 90.8% as malignant and 9.2% as benign. Biopsy confirmed malignancy in 92.3% of the cases. Taking histopathology as the gold standard, FNAC demonstrated a sensitivity of 98.33%, specificity of 100%, positive predictive value(PPV) of 100%, and negative predictive value (NPV) of 83.33%. Concordance between FNAC and histopathological subtyping was 98.46%. Adenocarcinoma was the most common subtype, followed by squamous cell carcinoma and small cell carcinoma. Smoking was the most common contributing factor associated with malignancy.

**Conclusion and implications:** Image-guided FNAC is an excellent diagnostic accuracy tool which possess higher level of concordance with biopsy in evaluating lung masses. It should be considered as the frontline diagnostic tool, especially in resource limited settings.

## INTRODUCTION

Lung cancer remains one of the most significant causes of cancer-related morbidity and mortality globally.^(1)^ According to the Global Cancer Observatory (GLOBOCAN) 2020, lung cancer ranks second in incidence and first in cancer-related deaths worldwide.^(1)^ In Nepal, it is the leading cancer among males and among the top five in females. Timely and accurate diagnosis plays a vital role in effective management and prognosis of the disease.^(2)^

Image-guided fine needle aspiration cytology(FNAC) has emerged as a valuable diagnostic tool for evaluating pulmonary lesions, particularly in cases where the tumor is inaccessible by bronchoscopy.^(3)^ It is minimally invasive, cost-effective, and offers rapid preliminary diagnosis with fewer complications compared to core biopsies or surgical procedures.^(4)^

FNAC can provide early cytological diagnosis and, when coupled with imaging techniques such as CT or ultrasound, its diagnostic yield improves significantly.^(4, 5)^ Despite its proven utility, FNAC is still underutilized in some resource-limited settings like Nepal, often due to lack of expertise or infrastructural challenges.

Previous studies have reported variable diagnostic accuracy of FNAC in lung malignancies often ranging between 85% to 97%, depending on the type of lesion and technique used.^(6)^ Several studies have confirmed the diagnostic accuracy of image-guided FNAC for lung lesions. A study by Mondal et al.^(7)^ reported a sensitivity of 95.2% and specificity of 98.3% in diagnosing lung cancers using FNAC. Another multicentric trial by Nasit et al.^(8)^ demonstrated that FNAC combined with radiologic guidance offers high concordance with histopathological subtypes.

The diagnostic yield of FNAC can vary based on lesion location, size, imaging modality, and operater experience. Studies have consistently found that Computed Tomography FNAC (CT-guided FNAC) improves yield in deep parenchymal or small lesions, while ultrasound guided FNAC is advantageous in peripheral and pleural based lesions.^(9)^

In terms of cytologic classification, FNAC enables differentiation between major histologic types of lung cancer such as adenocarcinoma, squamous cell carcinoma, and small cell carcinoma. The World Health Organization (WHO) classification of lung tumors, revised in 2015 and updated in 2021, emphasizes the utility of morphology and immunocytochemistry for subtyping lung tumors-a principle applicable to FNAC smears.^(10)^

Despite these global advances, literature from Nepal is limited. Only a few institutional studies have evaluated the use of FNAC in lung tumor diagnosis. What is the diagnostic accuracy and feasibility of FNAC in evaluation of lung tumors in a tertiary care setting in Nepal? This research study directly address the identified gap between limited national data and need for standardization of FNAC in lung tumor diagnosis. These studies highlighted high accuracy and feasibility of FNAC but also emphasized the need for standardization and more widespread adoption.^(11)^

This review supports the current study’s rationale by consolidating global and regional evidence that image-guided FNAC is valid, safe and effective method for diagnosing lung tumors. The findings from this study would contribute to the growing body of evidence supporting its use, particularly in developing countries.

## MATERIALS AND METHODS

This study was a hospital-based cross-sectional study conducted at the Department of Pathology, Chitwan Medical College Teaching Hospital, Bharatpur, Nepal, over a period of eighteen months, from April 2023 to September 2024. Approval for the study was obtained from the Institutional Review Committee (IRC) of Chitwan Medical College (CMC-IRC Ref No: 2078/79/103). The study included 65 patients during the period of study who presented with a lung mass detected on imaging and underwent image-guided FNAC followed by biopsy at the same anatomical site. Patients with coagulopathies or poor general condition unsuitable for FNAC were excluded.

FNAC was performed using ultrasound or CT guidance, depending on the location of the lesion. A 21-23 gauge needle attached to a 10ml syringe was used for aspiration. Multiple passes were made as needed to obtain adequate material. Smears were air-dried and stained with May-Grunwald-Giemsa (MGG), while others were fixed in 95% for Papanicolaou staining. Smears were interpreted under light microscopy. Cytological categorization included: Adenocarcinoma, Squamous cell carcinoma, Small cell carcinoma, Other malignancies, and Benign/Inflammatory lesions. Histopathological evaluation included biopsy samples that were fixed in 10% buffered formalin, processed, embedded in paraffin, sectioned, and stained with hematoxylin and eosin (H and E). ^(12)^ Diagnosis were made following WHO 2021 classification of lung tumors.^(10)^

Results from FNAC were compared with final histopathological diagnosis. Only unequivocal diagnosis were considered positive. Diagnostic accuracy was calculated using standard statistical parameters

- Sensitivity = TP/(TP + FN)
- Specificity = TN/(TN + FP)
- Positive Predictive Value (PPV) = TP/ (TP + FP)
- Negative Predictive Value (NPV) = TN/ (TN + FN)

Concordance rate between FNAC subtype and histopathology was also evaluated. Data were compiled and analyzed using Microsoft Excel and SPSS version 20.0.

## RESULTS

The age range was 20 to 88 years, with the majority in the 71-80 year age groups. Female predominated with a male-to-female ratio of 5:8. The youngest patient was 20 years old female, while the oldest one was 88 years old male. (Table 1)

**Table 1:** Demographic profile of patients with lung lesions (n=65)

| Variable | Category | Number of patients(n) | Percentage(%) |
| --- | --- | --- | --- |
| Age(years) | 20-30 | 2 | 3.1 |
|  | 31-40 | 0 | 0 |
|  | 41-50 | 3 | 4.6 |
|  | 51-60 | 12 | 18.5 |
|  | 61-70 | 21 | 32.3 |
|  | 71-80 | 22 | 33.8 |
|  | 81-90 | 5 | 7.7 |
| Sex | Male | 25 | 38.5 |
|  | Female | 40 | 61.5 |

Among all the patients, 73.8% of the cases gave history of smoking. Cough accounted for 55.4% of the cases. Cough was a common symptom of lung cancer, so more than half of the patients experiencing this symptom aligns with its clinical presentation. Shortness of breath was reported as 44.6% of the cases. Others signs and symptoms were Hemoptysis, Chest pain and fever. 36.9% of the cases complained of extra symptoms like hemoptysis, chest pain and/or fever. (Table 2)

**Table 2:** Clinical history of cases (n=65)

| Variables | Frequency (%) |
| --- | --- |
| **Smoking habit** |  |
| Yes | 73.8 |
| No | 26.2 |
| **Cough** |  |
| Yes | 55.4 |
| No | 44.6 |
| **Shortness of breath** |  |
| Yes | 44.6 |
| No | 55.4 |
| **Others<sup>##</sup>** |  |
| Yes | 36.9 |
| No | 63.1 |

*## denotes Hemoptysis, Chest pain, Fever*

Squamous Cell Carcinoma was the most common type of tumor across all age groups, particularly in middle aged (40-64 yrs) and elder aged (65 and above) groups. Adenocarcinoma was more frequently seen in the middle-aged group, but also appeared in other age groups. Neuroendocrine Tumors and Small Cell Carcinoma are less common but present in both FNAC and histopathology reports. Suspicious for malignancy appears across various age groups but is particularly noted in FNAC findings for middle aged and elder aged groups.

Table 3 shows the distribution of malignant and benign findings identified through image-guided FNAC (Fine Needle Aspiration Cytology) in the study of lung tumor. Adenocarcinoma was the most frequently diagnosed malignant tumor, accounting for 40.7% (24 cases) of the total malignant cases. Squamous cell carcinoma was identified in 22% (13 cases) of the malignancy. Small cell carcinoma was diagnosed in 15.3% (9 cases). Suspicious for malignancy accounted for 20.3% (12 cases). A single case (1.7%) of a neuroendocrine tumor was recorded. Remaining 9.2% (6 cases) of total cases were found to be benign.

**Table 3:** Diagnosis by image guided FNAC (n=65)

| Variables | Frequency | Percentage (%) |
| --- | --- | --- |
| **FNAC findings** |  |  |
| Malignant | 59 | 90.8 |
| Benign | 6 | 9.2 |
| **If malignant, then types (n=59)** |  |  |
| Adenocarcinoma | 24 | 40.7 |
| Squamous cell carcinoma | 13 | 22 |
| Small cell carcinoma | 9 | 15.3 |
| Suspicious for malignancy | 12 | 20.3 |
| Neuroendocrine tumor | 1 | 1.7 |

The table 4 presents the distribution of benign and malignant findings through image guided biopsy in a study population of 65 patients. The most frequently diagnosed malignant tumor was adenocarcinoma, representing 43.3% of the malignant cases (26 out of 60). Squamous cell carcinoma was the second most common, accounting for 26.7% of cases (16 out of 60), followed by small cell carcinoma, observed in 15% of cases (9 out of 60). Other less frequent malignancies included were Poorly differentiated carcinoma presenting in 6.6% of cases (4 out of 60), Non-small cell carcinoma in 5% (3 out of 60), Neuroendocrine tumor in 1.7% (1 out of 60) and Malignant mesothelioma in 1.7% (1 out of 60). Rest of 7.7% (5 out of 65 total cases) were benign.

**Table 4:**
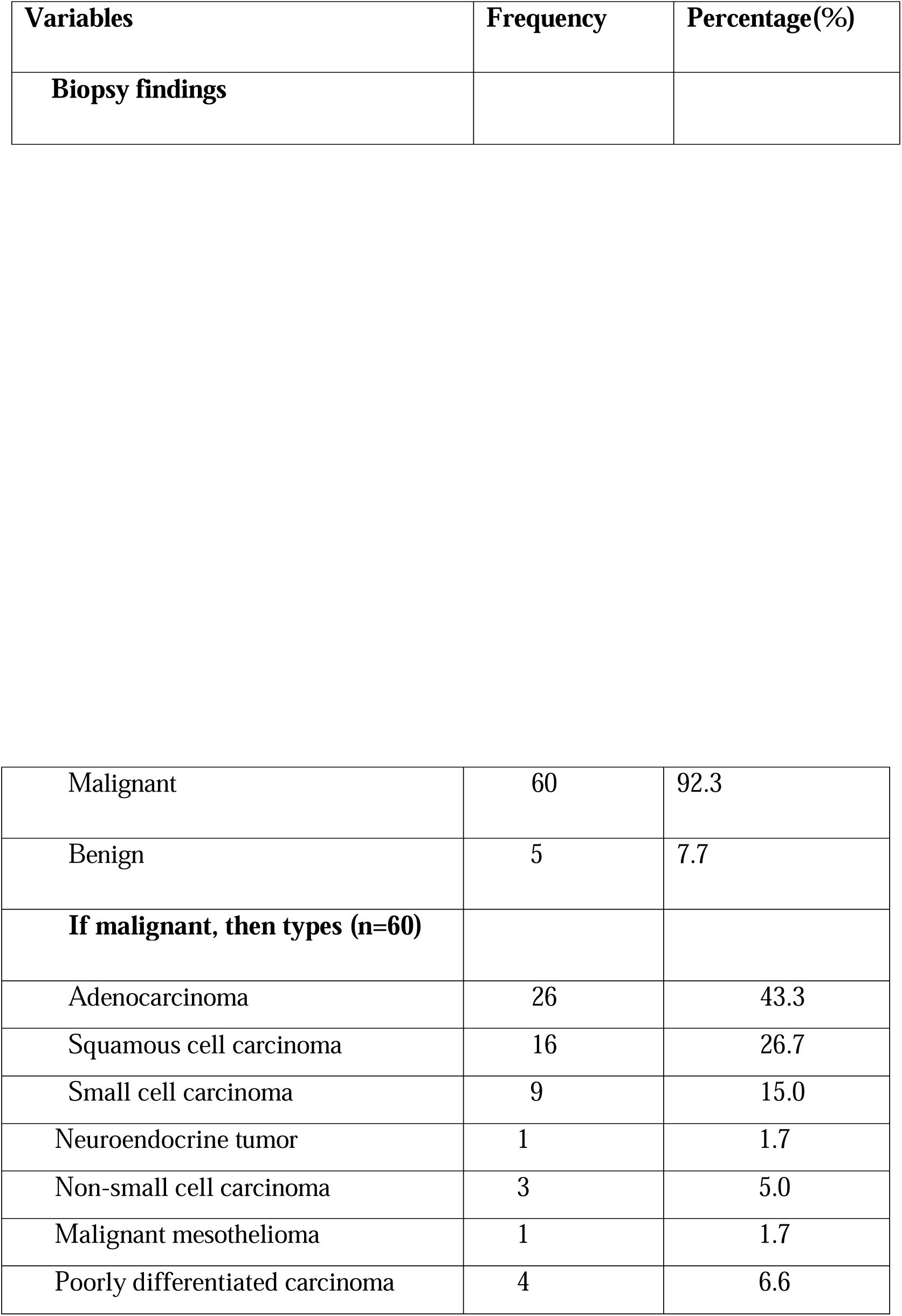
Diagnosis by image guided biopsy (n=65)

The table 5 indicates FNAC findings correlating with biopsy as gold standard tool. There were 59(90.8%) true positive cases that were confirmed as malignant type by FNAC and biopsy. Only a single case (1.5%) regarded as false negative case showed benign characteristic in FNAC but diagnosed as poorly differentiated carcinoma, a malignant tumor on histopathology. Remaining 5 (7.7%) true negative cases of lung nodule were diagnosed with benign characteristics, both on FNAC and biopsy.

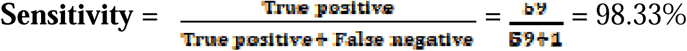

**Table 5:** Accuracy of image guided FNAC with biopsy finding (n=65)

| FNAC finding | Biopsy finding |  | Total |
| --- | --- | --- | --- |
|  | Positive (%) | Negative (%) |  |
| **Positive** | 59 (90.8%) | 0 | 59 (90.8%) |
| **Negative** | 1 (1.5%) | 5 (7.7%) | 6 (9.2%) |
| **Total** | 60 (92.3%) | 5 (7.7%) | 65 (100%) |

This indicates FNAC correctly identified 98.33% of the actual positive cases.

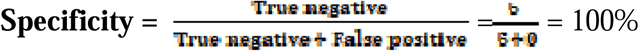

This high specificity indicates that when FNAC identifies a negative finding, it is correct 100% of the time.

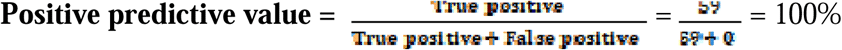

This means that when FNAC indicates a positive finding, there is 100% chance that it corresponds to a true malignancy.

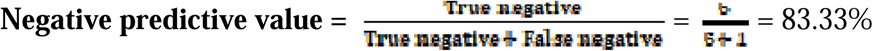

This indicates that when FNAC yields a negative result, there is a 83.33% probability that the patient does not have malignancy.

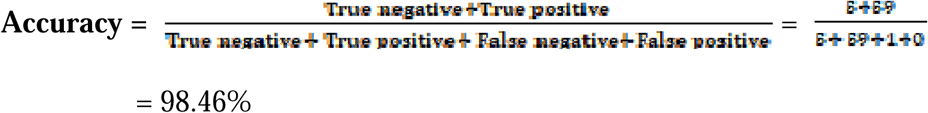

This indicates that out of all 65 patients undergoing FNAC, 98.46% were correctly diagnosed either as having the tumor(true positive) or not having it (true negative).

**Photograph 1:**
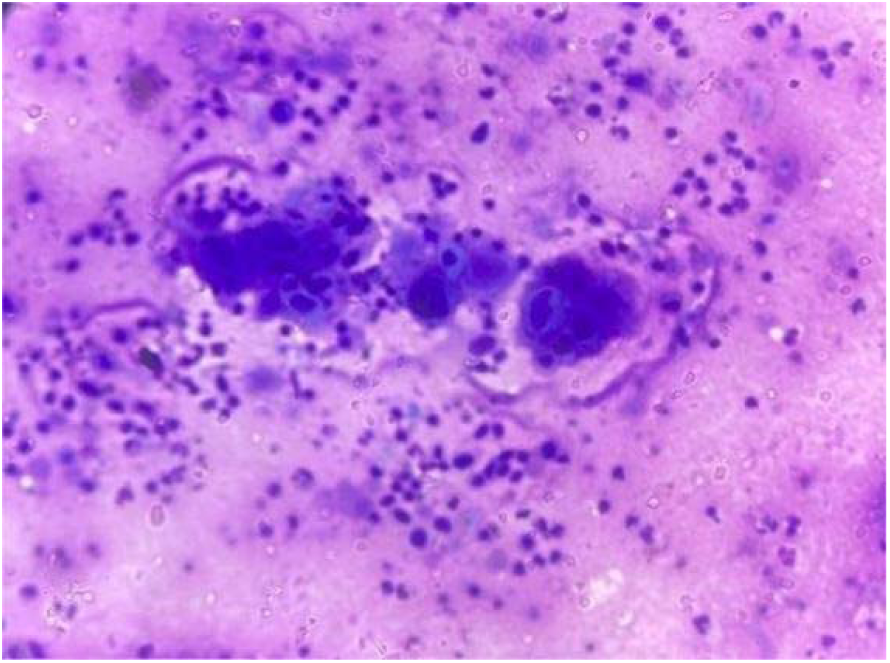
Giemsa stain x400: FNAC smear showing Squamous Cell Carcinoma (SCC)

**Photograph 2:**
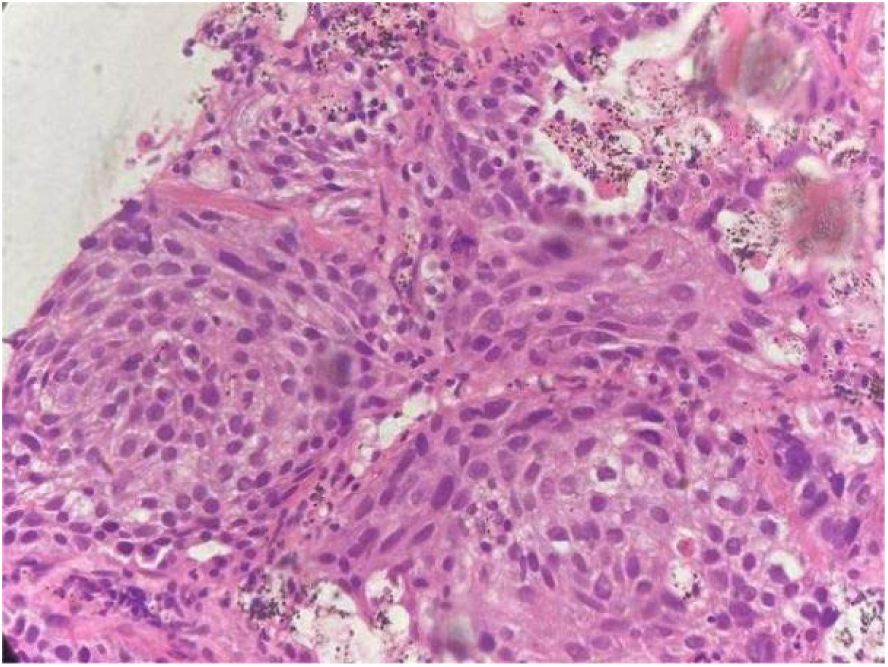
H and E stain x400: Section showing Squamous Cell Carcinoma (SCC)

**Photograph 3:**
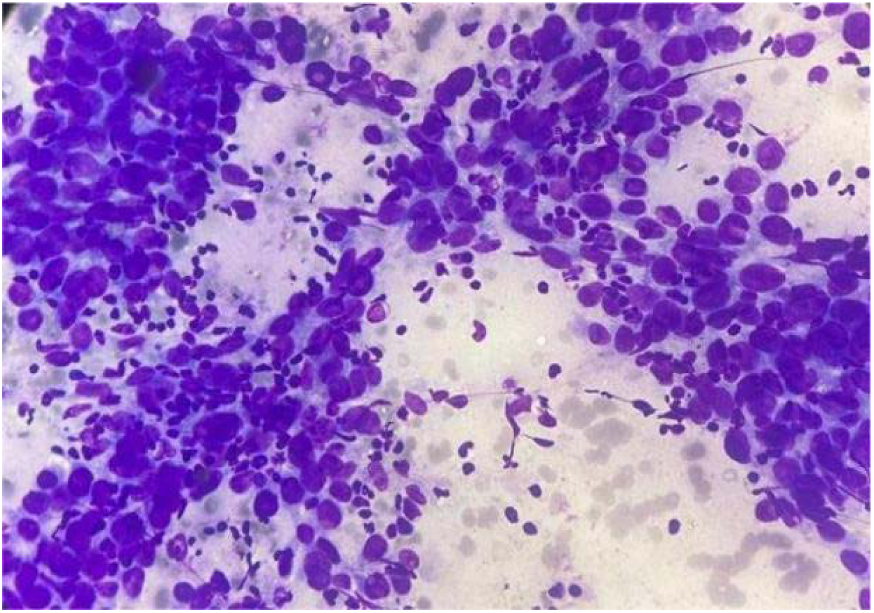
Giemsa stain x400: Smear showing Adenocarcinoma.

**Photograph 4:**
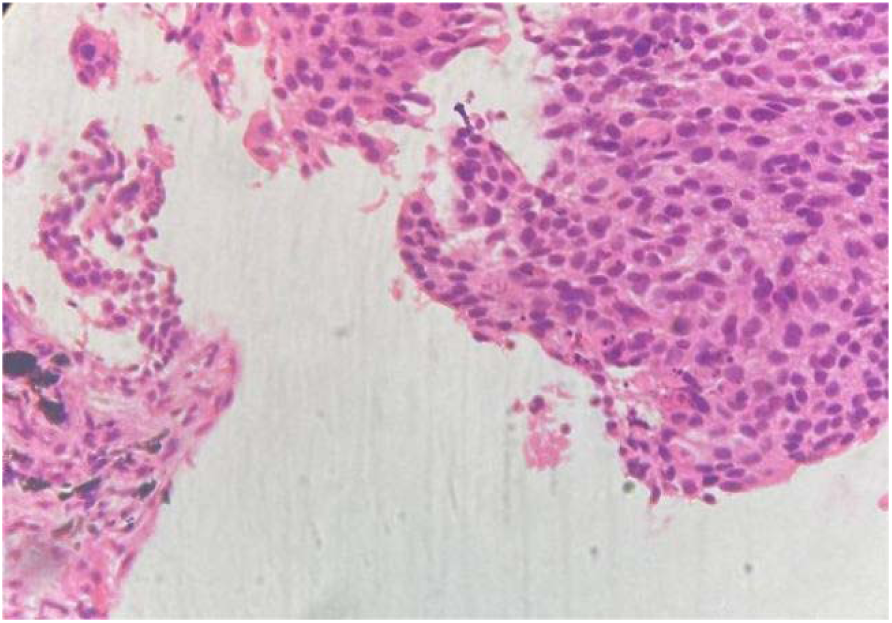
H and E stain x400: Section showing Non-small Cell Carcinoma.

**Photograph 5:**
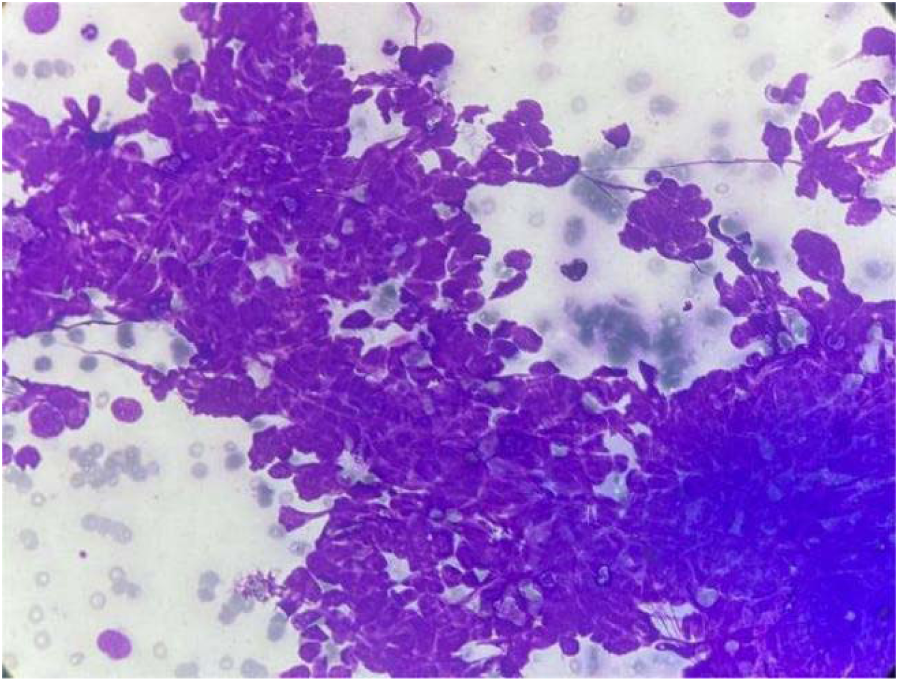
Giemsa stain x400: Smear showing Small Cell Carcinoma.

**Photograph 6:**
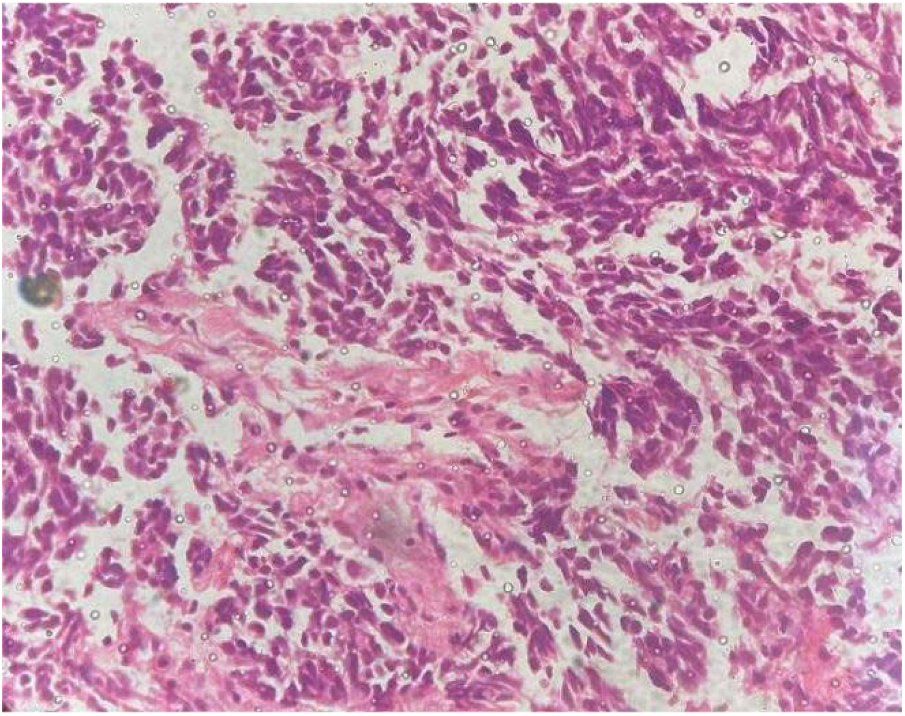
H and E stain x400: Section showing Small Cell Carcinoma.

**Photograph 7:**
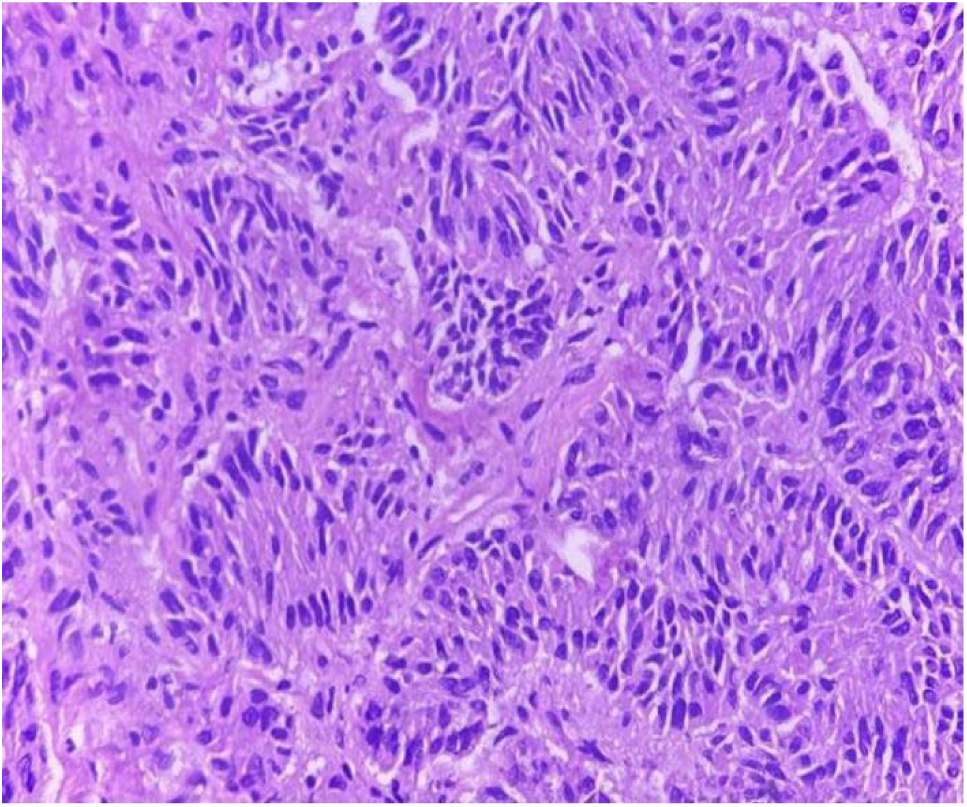
H and E stain x400: Section showing Carcinoid tumor.

## DISCUSSION

Our study demonstrates the significant clinical utility of Fine Needle Aspiration Cytology (FNAC) in the diagnosis of lung tumors. A high proportion (73.8%) of patients had a history of smoking, a well established risk factor for lung malignancy. Respiratory symptoms were common, with cough (55.4%) and shortness of breath (44.6%) being the most most frequently reported, followed by chest pain, fever, and hemoptysis (36.9%). These findings are consistent with previous literature indicating that such symptoms often prompt further diagnostic investigation, including FNAC.^(13)^

FNAC proved to be a highly effective diagnostic modality in our study. Among the malignancies diagnosed, adenocarcinoma was the most prevalent (40.7%), followed by squamous cell carcinoma (22%) and small cell carcinoma (15.3%).^(14, 15)^ Additionally, 20.3% were classified as “Suspicious for malignancy” indicating that while FNAC is highly accurate, some cases require further testing or biopsy. Rare tumors, such as neuroendocrine tumors, were identified in 1.7%, and benign lesions accounted for 9.2%, confirming FNAC’s ability to differentiate between malignant and non-malignant lesions, thereby reducing the need for more invasive procedures. Overall, FNAC demonstrates strong diagnostic accuracy but may require additional methods in inconclusive cases, while its minimally invasive nature and high accuracy for common malignancies underscore its clinical value.

Comparative studies support these findings. Mondal et al. (2013)^(7)^ and Maman et al. (2024)^(16)^ reported image-guided biopsy results with adenocarcinoma as the most common malignancy (43.3%), followed by squamous cell carcinoma (26.7%), small cell carcinoma (15%), and poorly differentiated carcinoma (6.6%), alongside rare tumors like neuroendocrine tumors and malignant mesothelioma. The FNAC study showed adenocarcinoma in 40.7% of cases, closely matching biopsy results, underscoring FNAC as a reliable, less invasive alternative. Squamous cell carcinoma detection by FNAC (22%) and biopsy (26.7%) were comparable, and small cell carcinoma detection was nearly identical (FNAC 15.3%, biopsy 15%). Poorly differentiated carcinoma was detected in 6.6% of biopsy cases but not explicitly in FNAC, reflecting biopsy’s higher histopathological detail. FNAC’s combination with molecular techniques or immunohistochemistry could enhance tumor differentiation, especially in the 20.3% “suspicious for malignancy” cases. Both FNAC and biopsy detected neuroendocrine tumors (1.7%), and malignant mesothelioma was reported in 1.7% of biospy cases. Benign lesions accounted for 7.7% in biopsy and 9.2% in FNAC, consistent with Biancosino et al. (2016)^(14)^ confirming FNAC’s reliablity in ruling out malignancy. FNAC offers a less invasive approach with diagnostic accuracy comparable to biopsy for common malignancies and can serve as an essential first-line tool, with biopsy reserved for suspicious or inconclusive cases.

In our study, FNAC demonstrated high diagnostic accuracy, with 90.8% true positives (59 cases accurately diagnosed as malignant), one false negative (1.5%) where FNAC missed poorly differentiated carcinoma, and 7.7% true negatives for benign lesions. These results align with literature, such as Dahlstrom et al. (2001)^(17)^, reporting FNAC sensitivity of 92% and specificity of 100%, noting occasional false negatives in poorly differentiated tumors. Abdulrahman et al. (2024)^(18)^ reported 86.9% sensitivity, with FNAC effective in distinguishing malignant from benign lesions but limited in subtle cytological features. Nizzoli et al. (2011)^(19)^ noted around 10% false negatives due to sampling errors or deep lesion access challenges. Our lower false-negative rate (1.5%) may reflect improved techniques or pathologist experience. FNAC’s minimally invasive and cost-effective nature makes it highly reliable, though sensitivity may vary with tumor morphology, size, and operator expertise. Overall, FNAC’s diagnostic performance in our study compares favorably with previous research, emphasizing its utility as an initial diagnostic tool while acknowledging the need for biopsy in ambiguous or clinically suspicious cases.

Sensitivity, specificity, and predictive values further support FNAC’s effectiveness. Sensitivity was 98.33%, correctly identifying nearly all true positives. Specificity was 100%, indicating all negative cases were truly negative. Positive Predictive Value (PPV) was 100%, meaning all positive FNAC results corresponded to actual malignancies. These values surpass average literature sensitivity for FNAC (85-95%), likely due to sample size, pathologist expertise, and imaging guidance. High specificity and PPV reflect FNAC’s robustness in avoiding false positives, allowing confident diagnosis of malignancy.^(20)^ However, replicability in larger, diverse populations may vary due to equipment, operator skill, and tumor accessibility. The negative predictive value (NPV) was 83.33%, indicating reasonable reliablity in ruling out malignancy, though around 16.67% of negative FNAC cases could still harbor malignancy, emphasizing follow-up or repeat biopsy in high-suspicion cases. Factors affecting NPV include prevalence, tumor location, size, and imaging guidance, with false negatives often due to sampling errors, necrosis, or cellular heterogeneity.^(21)^

In our cohort of 65 patients, FNAC accurately diagnosed 98.46%, combining true positives and true negatives. The high diagnostic accuracy aligns with, and in some cases surpasses, other studies: Qureshi et al. (2024)^(22)^ reported >95% accuracy using ultrasound-guided FNAC, and Ekheiwish et al. (2010)^(20)^ found 84-95% accuracy in meta-analysis. Banik et al. (2023)^(23)^ reported 90% accuracy for centrally located lesions using transbronchial FNAC. FNAC generally yields higher accuracy for central masses compared to peripheral nodules, with imaging guidance (ultrasound, endobronchial ultrasound) further enhancing precision and diagnostic yield. The high accuracy in our study reinforces FNAC as a minimally invasive, reliable diagnostic method, especially when integrated with imaging guidance.

## CONCLUSION

In conclusion, our study demonstrates FNAC’s high sensitivity (98.33%), specificity (100%), PPV (100%), and NPV (83.33%) for lung tumor diagnosis, with overall diagnostic accuracy of 98.46%. FNAC effectively detects common malignancies such as adenocarcinoma, squamous cell carcinoma, and small cell carcinoma, while also identifying rare tumors. Though false negatives, especially in poorly differentiated carcinomas, remain a limitation, FNAC offers a less invaisve, cost-effective, and accurate diagnostic approach. When combined with imaging guidance and supplemented with biopsy or molecular studies in ambiguous cases, FNAC provides a reliable first-line tool for the early detection and diagnosis of lung tumors, supporting clinical decision-making and optimizing patient care.

## IMPLICATIONS

1. High accuracy of FNAC in lung tumor evaluation supports its use as a reliable, minimally invasive and cost-effective diagnostic tool, facilitating early diagnosis and timely initiation of therapy.
2. The results highlight the need to establish standardized FNAC protocols and quality assurance systems, expand cytology services to peripheral hospitals, and integrate FNAC data into national cancer registry programs to strengthen cancer control strategies in Nepal.
3. For future research, this study provides a foundation for multicenter validation, incorporation of ancillary techniques such as cell block and immunocytochemistry, and exploration of molecular or digital cytology applications, which can further enhance diagnostic precision and promote personalized management of lung cancer.

## LIMITATIONS

1. The results of the present study were based on a small population sample size which limits their application to the larger population sample size.
2. This study was conducted in a single center, which may limit the applicability of the results to other clinical settings where different techniques or expertise levels may be present.
3. The procedure’s sensitivity and negative predictive value could be influenced by lesion size and location. Small nodules are more challenging to access and may result in a higher rate of false-negative results.

## Data Availability

All data produced in the present study are available upon reasonable request to the authors

## Declaration of interests

⍰The authors declare that they have no known competing financial interests or personal relationships that could have appeared to influence the work reported in this paper.

## Author biography

Dr. Bishwobandhu Bhandari is a Pathologist from Nepal specializing in histopathology, cytopathology and hematopathology. His research interests include in the field of histopathology, cytopathology and hematopathology along with molecular diagnostics. He is currently involved in clinical fellowship program in hematopathology department at Tribhuvan University Teaching Hospital, Institute of Medicine, Kathmandu, Nepal. He has participated in academic research and medical education activities in pathology at Nepal.

## Notes

### Competing Interest Statement

The authors have declared no competing interest.

### Author Declarations

Approval for the study was obtained from the Institutional Review Committee (IRC) of Chitwan Medical College (CMC-IRC Ref No: 2078/79/103).

## REFERENCES

1. Sharma R. Mapping of global, regional, and national incidence, mortality and mortality to incidence ratio of lung cancer in 2020 and 2050. Int J Clin Oncol. 2022;27(4):665–675. 10.1007/s10147-021-02108-2.

2. Shilpakar R, Paudel BD, Sharma R, Silwal SR, Sapkota R, Shrestha P, et al. Lung cancer in Nepal. J Thorac Oncol. 2022;17(1):2229. 10.1016/j.jtho.2021.09.021.

3. Martin HE, Ellis EB. Biopsy by needle puncture and aspiration. Ann Surg. 1930;92(2):169–181. 10.1097/00000658-193008000-00001.

4. Ahmad M, Afzal S, Saeed W, Mubarik A, Saleem N, Khan SA, et al. Efficacy of bronchial wash cytology and its correlation with biopsy in lung tumours. J Pak Med Assoc. 2004;54(1):13–16.

5. Mullan CP, Kelly BE, Ellis PK, Hughes S, Anderson N, Mc Cluggage WG. CT-guided fine needle aspiration of lung nodules: effect on outcome of using coaxial technique and immediate cytological evaluation. Ulster Med J. 2004 May;73(1):32–36

6. Lin R, Che G. Validation of the Mandarin Chinese version of the Leicester Cough Questionnaire in non-small cell lung cancer patients after surgery. Thorac Cancer. 2018;9(4):486–90. doi:10.1111/1759-7714.12602

7. Mondal SK, Nag D, Das R, Mandal PK, Biswas PK, Osta M. Computed tomogram guided fine-needle aspiration cytology of lung mass with histological correlation: A study in Eastern India. South Asian J Cancer. 2013;2(1):14–8. doi:10.4103/2278-330X.105881

8. Nasit JG, Parikh PK, Shah MB, Davara K, Sharma P. Diagnostic utility of fine needle aspiration cytology in lung cancer: A study in Eastern India. Indian J Pathol Microbiol. 2013;30:179–83. [DOI not available]

9. Wallace MJ, Krishnamurthy S, Broemeling LD, Gupta S, Ahrar K, Morello FA Jr, et al. CT guided percutaneous fine needle aspiration biopsy of small(≤1cm) pulmonarylesions. Radiology.2002;225(3):823–8.doi:10.1148/radiol.2253011465

10. Singh G, Singh A, Dave R. An update on WHO classification of thoracic tumours 2021-newly described entities and terminologies. J Clin Diagn Res.2023;17(6):EE01–5. doi:10.7860/JCDR/2023/62583.18076

11. Ranabhat S, Poudel S, Pun G, Awale A. Role of image guided fine needle aspiration cytology of lung lesions in patients visiting Gandaki Medical College. J Pathol Nepal. 2022;12(2):1943–9. doi:10.3126/jpn.v12i2.51747

12. Bancroft JD, Gamble M. Theory and practice of histological techniques. 6th ed. London: Elsevier Health Sciences; 2008. [Book—no DOI]

13. Pial SA. A cross-sectional study on risk factors, symptoms, treatment and management strategies of lung cancer in a tertiary care cancer hospital of Bangladesh [dissertation]. 2018. [Thesis—no DOI]

14. Biancosino C, Krüger M, Vollmer E, Welker L. Intraoperative fine needle aspirations— diagnosis and typing of lung cancer in small biopsies: challenges and limitations. Diagn Pathol. 2016;11(1):59. doi:10.1186/s13000-016-0505-3

15. Rudin CM, Brambilla E, Faivre-Finn C, Sage J. Small-cell lung cancer. Nat Rev Dis Primers. 2021;7(1):3. doi:10.1038/s41572-020-00235-0

16. Maman A, Çiğdem S, Kaya İ, Demirtaş R, Ceylan O, Özmen S. Diagnostic value of FDG PET-CT in differentiating lung adenocarcinoma from squamous cell carcinoma. EJNMMI Rep. 2024;8(1):1. doi:10.1186/s41824-023-00185-7

17. Dahlstrom JE, Langdale-Smith GM, James DT. Fine needle aspiration cytology of pulmonary lesions: a reliable diagnostic test. Pathology. 2001;33(1):13–6. doi:10.1080/00313020120033470

18. Abdulrahman AN, Salman HH. Evaluate the reliability of fine-needle aspiration cytology (FNAC) in determining the accurate diagnosis of thoracic lesions. Eur J Mod Med Pract. 2024;4(4):74–8. [DOI not available]

19. Nizzoli R, Tiseo M, Gelsomino F, Bartolotti M, Majori M, Ferrari L, et al. Accuracy of fine needle aspiration cytology in the pathological typing of non-small cell lung cancer. J Thorac Oncol. 2011;6(3):489–93. doi:10.1097/JTO.0b013e31820c4c3d

20. Ekheiwish HS, Rasool HA. The role of ultrasound-guided lung FNAC exam in the diagnosis of bronchogenic carcinoma. Med J Babylon. 2010;7(4):3. [DOI not available]

21. Mukherjee S, Bandyopadhyay G, Bhattacharya A, Ghosh R, Barui G, Karmakar R. Computed tomography-guided fine needle aspiration cytology of solitary pulmonary nodules suspected to be bronchogenic carcinoma: Experience of a general hospital. J Cytol. 2010;27(1):8–11. doi:10.4103/0970-9371.66695

22. Qureshi AR, Mumtaz B, Akhtar Z, Ashraf Z, Sajid M, Muhammad A. Ultrasound guided fine needle aspiration cytology: An effective diagnostic tool in pulmonary medicine. Pak Armed Forces Med J. 2024;74(3):626–30. [DOI not available]

23. Banik T, Bhattacharyya R, Basu N, Majee SS. Role of FNAC in early detection and diagnosis of lung lesions with histo-radiological correlation and clinical insights in a tertiary care centre. J Med Sci Health. 2023;9(1):92–8. doi:10.46347/jmsh.2023.v09i01.016

